# Predicted CTL responses from pressured epitopes in SARS-CoV-2 correlate with COVID-19 severity

**DOI:** 10.1101/2021.12.06.21267084

**Authors:** Vishal Rao, Ushashi Banerjee, Narmada Sambaturu, Sneha Chunchanur, R Ambica, Nagasuma Chandra

## Abstract

Heterogeneity in susceptibility among individuals to COVID-19 has been evident through the pandemic worldwide. Protective cytotoxic T lymphocyte (CTL) responses generated against pathogens in certain individuals are known to impose selection pressure on the pathogen, thus driving emergence of new variants. In this study, we focus on the role played by host genetic heterogeneity in terms of HLA-genotypes in determining differential COVID-19 severity in patients and dictating mechanisms of immune evasion adopted by SARS-CoV-2 due to the imposed immune pressure at global and cohort levels. We use bioinformatic tools for CTL epitope prediction to identify epitopes under immune pressure. Using HLA-genotype data of COVID-19 patients from a local cohort, we observe that asymptomatic individuals recognize a larger number of pressured epitopes which could facilitate emergence of mutations at these epitopic regions to overcome the protectivity they offer to the host. Based on the severity of COVID-19, we also identify HLA-alleles and epitopes that offer higher protectivity against severe disease in infected individuals. Finally, we shortlist a set of pressured and protective epitopes that represent regions in the viral proteome that are under higher immune pressure across SARS-CoV-2 variants due to the protectivity they offer. Identification of such epitopes could potentially aid in prediction of indigenous variants of SARS-CoV-2 and other pathogens, defined by the distribution of HLA-genotypes among members of a population.

## Introduction

Severe Acute Respiratory Syndrome Corona-Virus 2 (SARS-CoV-2) which is responsible for the Coronavirus Disease 2019 (COVID-19) pandemic, has infected over 259 million people around the world as of 26^th^ November, 2021^1^. Despite large-scale vaccination drives, the pandemic continues to spread owing to newly emerging variants of concern. Infection with SARS-CoV-2 presents a wide range of clinical outcomes ranging from asymptomatic, mild and moderate disease states to severe and critical states associated with severe pneumonia, Acute Respiratory Distress Syndrome (ARDS)^2^ as well as lymphopenia^3^. The immune mechanisms responsible for containing the viral infection has been shown to be dysfunctional in case of severe COVID-19 with sustained cytokine production leading to a hyper-inflamed state^4^. One of the major arms of the immune system implicated in combating viral infections is the Cytotoxic T Lymphocyte (CTL) or CD8+ T cell response, which exhibits a functionally exhausted phenotype in severe COVID-19 cases^5^. However, significant heterogeneity has been observed in the immunopathology across patients suffering from severe COVID-19 recommending the use of distinct therapeutic interventions for patients with different immunotypes ^6^.

Genetic heterogeneity across humans is known to majorly influence severity as well as immunopathology of a disease. The Human Leukocyte Antigen (HLA) genotype is one of the host factors which exhibits high sequence variation across individuals. The HLA Class I genotype, comprised of 3 sets of HLA alleles – A, B and C, can directly influence the CTL response which makes individuals with certain HLA genotypes more susceptible to certain diseases. Previous studies have identified many HLA Class-I alleles which are significantly associated with susceptibility to severe COVID-19^7^. Several advances in identifying immunodominant T cell epitopes have facilitated the quantification of T cell responses through prediction of binding affinities of HLA alleles to pathogenic epitopes using various bioinformatic techniques and machine-learning algorithms^8^. These epitope prediction tools have been used to determine susceptibilities of HLA alleles to severe COVID-19 based on epitope recognition^9^ and to identify peptide-based vaccine targets based on high population coverage of epitopes^10,11^,12. In addition, structural prediction of epitopes has also provided insights regarding constrained regions of the viral proteome that are robust to mutations^13,14^.

Host immune responses typically impose selection pressure on the pathogen, driving emergence of mutated variants that are capable of escaping the antiviral response. Among the variants of SARS-CoV-2, the Delta variant (B.1.617.2) has dominated over other variants during the intense COVID-19 wave in India in April-May 2021, due to its higher transmissibility and partial evasion of the host immune responses^15^. Previous studies have suggested the possibility of T cell immune escape in SARS-CoV-2 variants^16,17^,19 as well as isolates of the Wuhan strain^18^ either restricted to certain HLA alleles^16,17^,19 or covering larger populations^18,20^. On the contrary, mutations arising in the emerging variants have been reported to facilitate escape, mainly from the neutralizing antibody response rather than the T cell responses which are relatively more conserved across variants^21,22^. An effective T-cell response is highly dependent on presentation of certain epitopes that show high affinity binding with host HLA molecules. Hence, in order to analyse and explain mutation-driven escape of viral variants from host T cell responses, it becomes necessary to identify the epitopes which offer protectivity to the host and are therefore under selection pressure.

In this study, we aim to ascertain the role played by host genetic heterogeneity in determining differential COVID-19 susceptibility and severity across SARS-CoV-2 variants of concern (VOCs) and interest (VOIs). Using protein sequence data of SARS-CoV-2 variants available on the NCBI Virus database^23^, we obtain CTL epitope sets for each variant using the IEDB epitope prediction tools^24^. We then compare these epitope sets across variants and shortlist epitopes which are under immune pressure across variants. Next, we compare recognition of pressured epitopes across individuals with varying COVID-19 severity. Individuals who are more susceptible to severe COVID-19 are expected to recognize a smaller repertoire of epitopes by virtue of their HLA genotype and hence, elicit a poor CTL response against the virus. Finally, in a cohort generated from Bangalore, India, we identify epitopes and HLA-alleles, which offer protection from severe COVID-19 by assigning protectivity scores based on their recognition and occurrence respectively.

## Results

### CTL epitope prediction

Putative CTL epitopes in the SARS-CoV-2 proteome were predicted using the well-established NetMHCPanBA method made available by IEDB. Epitope predictions were carried out for the top 10 abundant proteins in the viral proteome, whose complete sequences were documented on the NCBI Virus database for (i) the 4 Variants of Concerns (VOCs) namely B.1.1.7 (Alpha), B.1.351 (Beta), P.1 (Gamma) and B.1.617.2 (Delta), (ii) 4 Variants of Interest (VOIs) namely B.1.525 (Eta), B.1.526 (Iota), B.1.617.1 (Kappa) and C.37 (Lambda) (as documented in the official WHO website as of August 2021) along with the Wuhan strain (A) and (iii) 2 non-virulent variants – B.1.526.1 and P.2. Predicted epitopes binding with their cognate HLA-alleles with IC50 < 50nM were classified as strong binding epitopes. Strong binding epitopes were predicted individually for all HLA Class I alleles documented across various ethnicities around the world in The Allele Frequency Net Database^25^. A total of 2022 strong binding epitopes exhibiting high affinity binding with 1299 HLA-alleles were identified (Supplementary File-1). This epitope pool was pruned to identify (a) epitopes conserved across all variants and (b) pressured epitopes – that are under negative selection pressure. A cohort from Bangalore consisting of 36 individuals with COVID-19 severity ranging from asymptomatic and mild to moderate and severe was analysed to obtain a theoretical measure of the CTL responses in each case which were then tested for any correlations with disease severity. CTL responses were predicted by identifying the HLA Class-I genotypes of each individual in the cohort, and identifying the pool of high-binding pressured epitopes, which was then consolidated in to a CTL response score. A schematic overview of the workflow is depicted in Figure 1.

**Figure 1:**
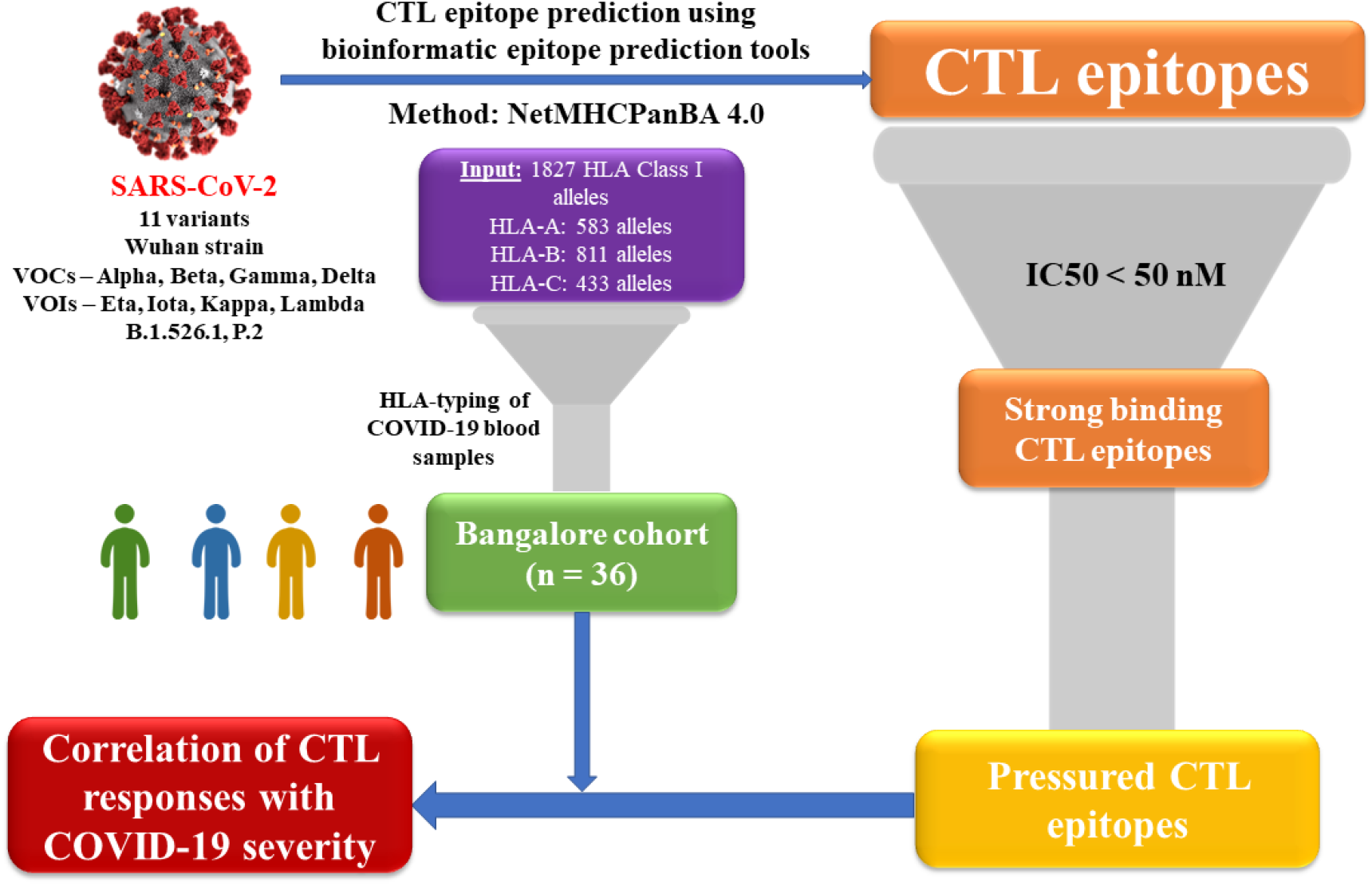
CTL Response Analysis Overview for SARS-CoV-2 Reference strain (Wuhan, Pangolin Lineage A) in comparison with SARS-CoV-2 Variants of Concern (VOCs), Variants of Interest (VOCs) and 2 non-virulent variants - B.1.526.1 and P.2. Using bioinformatic epitope prediction tools, CTL epitopes were predicted for the 10 SARS-CoV-2 proteins using the NetMHCPanBA method^28^. A total of 1827 HLA Class I alleles were used to predict binding affinities with SARS-CoV-2 epitopes. The initial epitope pool thus obtained was filtered based on peptide-HLA binding affinity (IC50 < 50 nM). From this shortlisted pool, epitopes under selection pressure across variants were identified and used for further analysis. COVID-19 blood samples from patients with a wide range of disease severity were collected and their HLA-genotypes were identified via HLA-typing. Finally, we performed correlation analysis between recognition of pressured epitopes and COVID-19 severity in patients.

### Immune pressure on CTL epitopes across SARS-CoV-2 variants

After shortlisting strong binding epitopes, we compared these epitope sets across variants of SARS-CoV-2. A pairwise estimate of difference in epitope sets as well as the protein sequence distance for each protein (Figure 2A, B) revealed higher variation in epitope sets compared to whole-protein sequence across the variants. This indicated that most proteins of SARS-CoV-2 are under immune pressure (Figure 2A, B, Supplementary Figure 1), except for Membrane protein and ORF6 (Supplementary Figure 1) which showed relatively higher conservation across variants. Immune pressure at the sequence level was evidenced by significant overlap between regions of high epitope density and higher variation, calculated as the Shannon entropy (Figure 2A, B).

**Figure 2:**
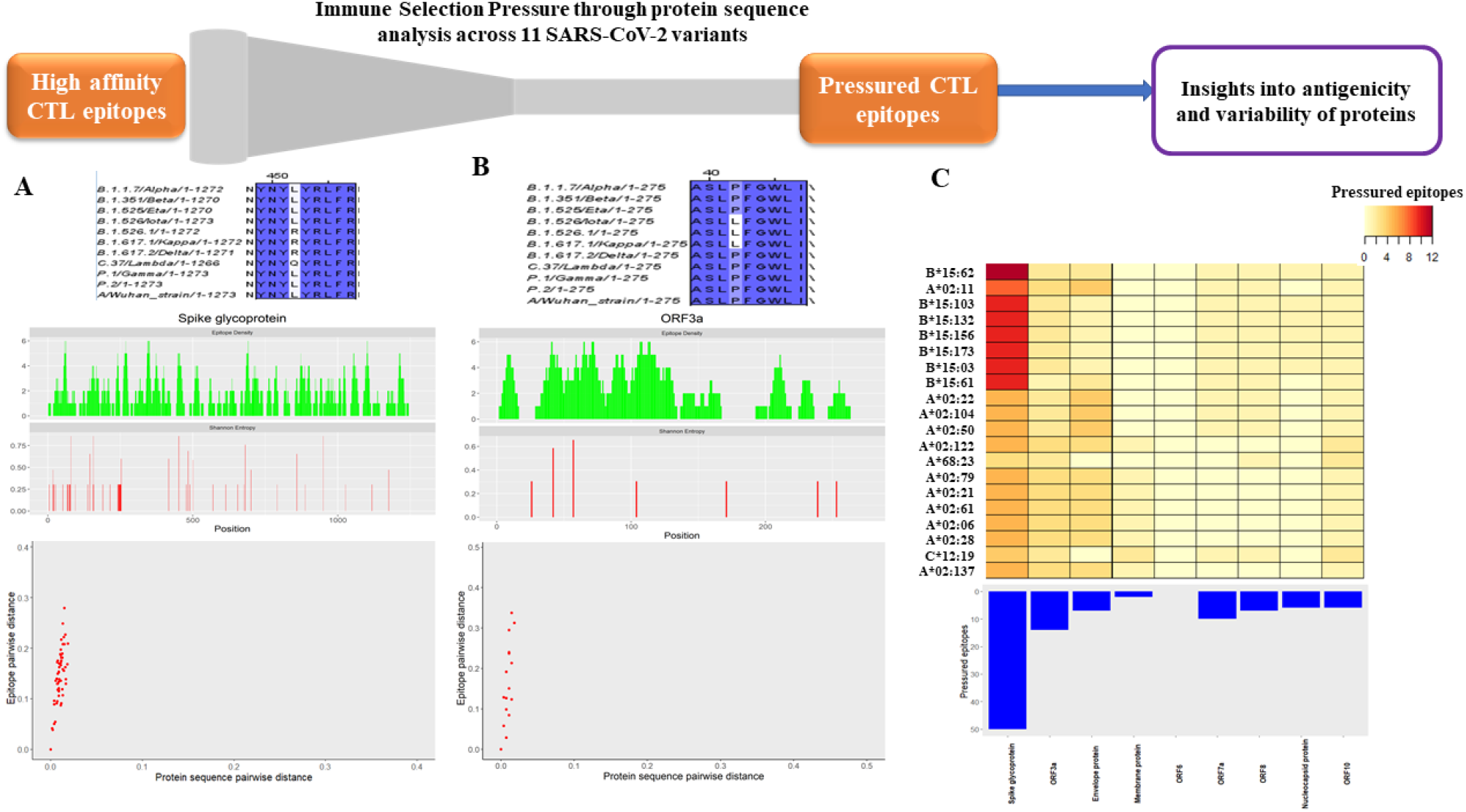
Analysis of CTL epitopes under immune pressure. Multiple sequence alignment of a representative epitope under immune pressure across the 11 variants of SARS-CoV-2 is depicted (top) for (A) Spike glycoprotein and (B) ORF3a protein. Symmetric difference between epitope sets for each pair of variants is plotted against the whole-protein sequence distance (bottom). The points in the scatter plot lie above the diagonal (y=x) for both proteins indicating higher selection pressure on epitopic regions. Sequence variation in the protein is computed as the Shannon entropy at each residue of the protein (middle - red). Distribution of epitopes within the protein is represented as the epitope density at each residue (middle - green). (C) Number of pressured epitopes in each protein of SARS-CoV-2, recognized by the HLA alleles (top 20 based on the number of epitopes recognized) are represented as a heat map (top). Contribution of each protein to the pressured epitope set is depicted by an inverted bar plot. ORF1ab is excluded from this plot since it is a polyprotein. Spike glycoprotein is seen to be the major contributor to the pressured epitope set.

Based on the observed variation across SARS-CoV-2 variants, from the pool of all strong binding epitopes, two subsets were selected: (a) epitopes which are conserved across all variants and (b) epitopes which are under selection pressure presumably due to host CTL responses and hence show sequence variation across variants. These pressured epitopes majorly belonged to the Spike glycoprotein (Figure 2C) while ORF6 lacked pressured epitopes as suggested by its high conservation across SARS-CoV-2 variants. We observed that HLA-B*15:62 followed by HLA-A*02:11 recognize the highest number of pressured epitopes among the HLA-Class I alleles considered. A ranked list of top-responding HLA-alleles is provided in Supplementary Table 1.

### Correlating predicted CTL responses with COVID-19 disease severity

Next, we tested if the pressured epitopes suggest the possibility of conferring protection against severe COVID-19 disease. Whole-blood samples from COVID-19 patients (n=36) with varying severity of the disease (Asymptomatic, Mild, Moderate and Severe) were collected from patients in Victoria Hospital, Bangalore, India and their HLA Class I genotypes were identified through their whole-blood RNA-seq data (Figure 3A). The predicted HLA genotypes for each of the samples in this Bangalore cohort are provided in Supplementary Table 2. The CTL response of an individual was calculated as the number of distinct epitopes recognized by the HLA alleles comprising the HLA genotype.

**Figure 3:**
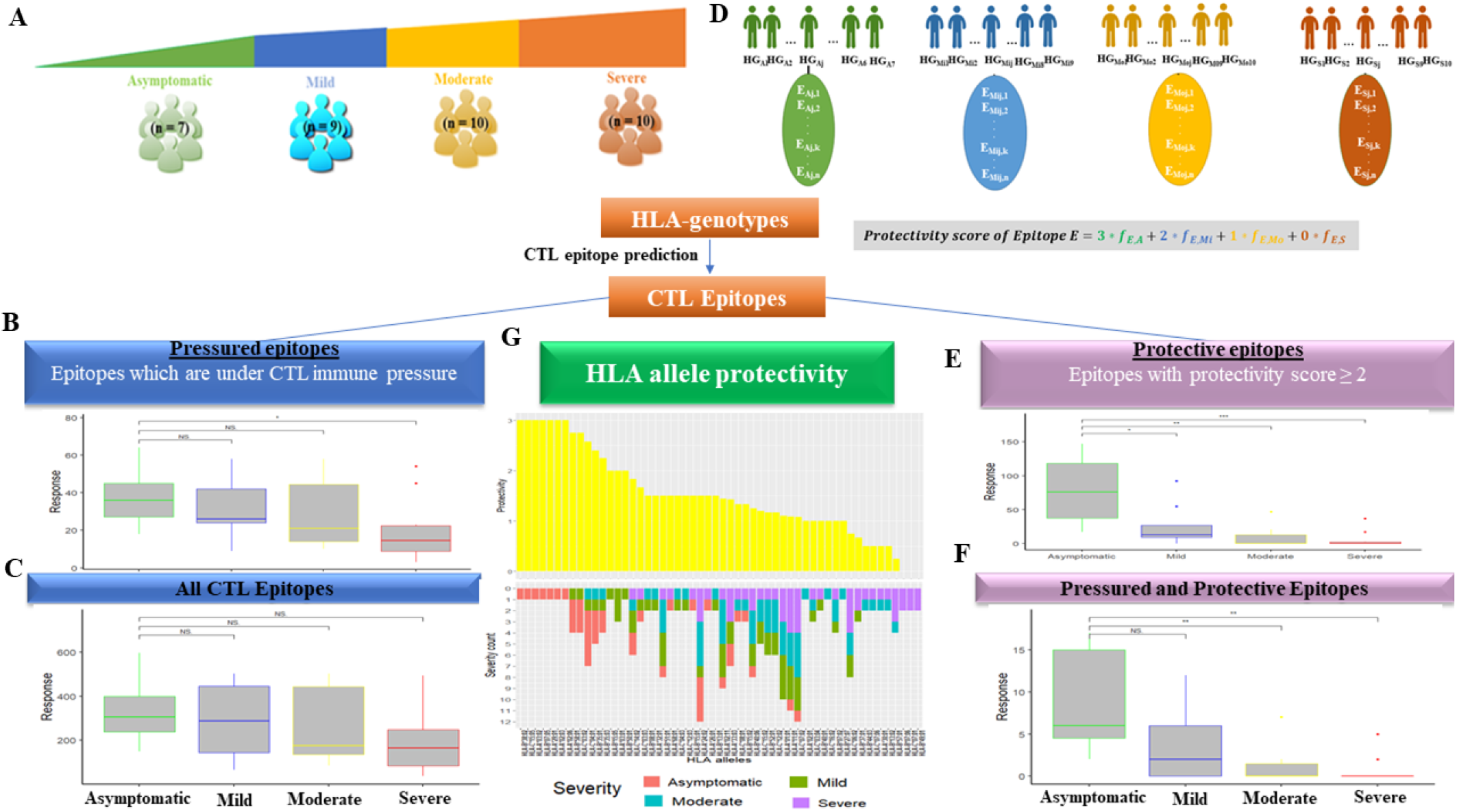
Correlation of predicted CTL responses with COVID-19 severity. (A) HLA-genotypes of COVID-19 patients from the Bangalore cohort, with 4 levels of disease severity - Asymptomatic, Mild, Moderate and Severe were identified using HLA-typing. CTL response measured in terms of the number of pressured epitopes (B) and (C) total number of epitopes recognized by individuals is plotted against the corresponding COVID-19 severity of the individual. Asymptomatic individuals recognize a significantly larger number of pressured epitopes as compared to those with severe COVID-19. (D) Schematic representation of computation of epitope protectivity score: Let [HG_Aj_]_1≤j≤7_, [HG_Mij_]_1≤j≤9_, [HG_Moj_]_1≤j≤10_ and [HG_Sj_]_1≤j≤10_ be the sets of individuals with asymptomatic, mild, moderate and severe COVID-19 respectively, represented by their HLA-genotypes. Each HLA genotype is associated with a corresponding set of distinct epitopes 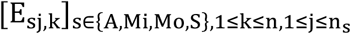 where n is the number of epitopes recognized by the HLA-alleles comprising the genotype and n_s_ is the number of individuals with disease severity s. Since epitope sets recognized by individuals with different disease severities overlap in many cases, the frequency of association of an epitope E with a disease severity DS, f_E,DS_ where DS ∈ {A, Mi, Mo, S} is calculated for each epitope. A protectivity value is then assigned for each disease state ranging from 3 for asymptomatic cases to 0 for severe cases. The protectivity score for an epitope is computed as the sum of the ‘protectivity value’ for each disease severity weighted by the frequency of association of the epitope with the particular disease severity (see Methods). (E) Protective CTL response calculated as the number of protective epitopes recognized by an individual is plotted against the corresponding COVID-19 severity of the individual. The number of protective epitopes recognized by asymptomatic individuals is significantly higher than those with mild, moderate or severe COVID-19. (F) The number of protective epitopes under immune pressure is plotted against disease severity of individuals recognizing these epitopes. Here, the number of pressured and protective epitopes recognized by asymptomatic individuals is significantly higher than those with moderate or severe COVID-19. (G) Protectivity scores of HLA alleles (top) based on the disease severity they associate with, along with the distribution of disease severity of patients expressing the allele (bottom) is shown. HLA alleles with higher protectivity are largely seen in Asymptomatic individuals while those with lower protectivity are observed in individuals with severe disease. Statistical significance for the bar plots was computed using the Wilcoxon paired signed-rank test (NS - Not Significant, * - p-value ≤ 0.05, ** - p-value ≤ 0.01, *** - p-value ≤ 0.001).

The resulting epitope pool of the entire cohort, referred to as the ‘all epitope’ pool, was further classified in 2 ways. Firstly, a set of locally pressured epitopes was shortlisted as a subset of the initially characterized globally pressured epitopes recognized by individuals within the Bangalore cohort. Secondly, epitopes which offered higher protectivity against severe COVID-19 were selected. Epitopes recognized by each HLA genotype within the cohort were assigned a protectivity score based on the severity of disease exhibited by the individual (Figure 3D). A pool of ‘protective’ epitopes was then constructed by shortlisting epitopes whose protectivity score crosses a set threshold corresponding to Asymptomatic and Mild disease states (see Methods).

We observed significantly higher recognition of pressured CTL epitopes by asymptomatic individuals as compared to severe cases (Figure 3B). In contrast, no significant difference was observed across disease states with the same analysis but using the ‘all epitope’ pool (Figure 3C). Our analysis shows that the overall CTL response decreases in the order of disease severity, with highest in asymptomatic and least in severe cases, suggesting that the HLA genotypes in asymptomatic individuals are the most protective in our cohort.

Next, we assigned a protectivity value for each disease severity, ranging from 0 for severe to 3 for asymptomatic disease states. We computed a protectivity score for each epitope as the sum of protectivity values of the 4 disease states, weighted by the frequency of occurrence of the epitope in individuals with the particular disease state. We observed a stronger correlation between the size of the protective epitope pool recognized and COVID-19 severity. The number of protective epitopes recognized by asymptomatic individuals was significantly higher than those recognized by mild, moderate and severe cases (Figure 3E). A pressured-protective epitope pool was then constructed by shortlisting pressured epitopes with high protectivity scores. A similar trend was observed where the size of the pressured-protective epitope pool was significantly higher in asymptomatic individuals as compared to those with moderate and severe disease (Figure 3F). A list of these pressured-protective epitopes is provided in Supplementary Table 3.

Next, we studied the association of individual HLA-alleles with disease severity in COVID-19 patients in the Bangalore cohort. We observed that many HLA-alleles mapped to multiple disease states in the cohort (Figure 3G). Similar to epitopes, a protectivity score was assigned to each HLA allele based on disease severity in the individual expressing the allele. HLA alleles A*02:03, A*02:06, A*03:02, A*29:01, B*07:05, B*38:02 and C*15:05 were seen to confer the highest protection while alleles B*07:06, B*49:01 and C*07:01 offered the least protection against severe disease. A list of HLA genotypes which offer highest protection against COVID-19 is provided in Supplementary Table 4. A gradient of disease states from completely asymptomatic in case of alleles with higher protectivity, to purely severe in case of alleles with least protectivity can be observed (Figure 3G) suggesting a major role of HLA Class-I mediated presentation of a selected epitope pool to CD8+ T cells in determining disease outcome.

## Discussion

Heterogeneity in HLA genotypes is known to influence differential disease susceptibilities among individuals. Such heterogeneity in HLA distribution is reflected in the wide range of disease outcomes observed in COVID-19, partially dictated by the T cell responses. Susceptibility is mainly attributed to occurrence of certain high-risk HLA alleles which bind poorly to SARS-CoV-2 epitopes thus resulting in a suboptimal CTL response^9.26^. However, analysis of CTL epitope sets in an individual at the HLA genotype level is needed to gain insights into possible mechanisms of immune escape adopted by the virus.

We performed CTL response analysis at the global level and within a cohort from Bangalore using HLA allele data. At the global level, we observe that CTL epitope sequences vary more than the rest of the proteome for most proteins, possibly indicating that SARS-CoV-2 variants are under T cell immune pressure. Based on this observation, we constructed a pressured epitope set comprising epitopes which are under immune pressure across the 11 SARS-CoV-2 variants considered. At the HLA genotype level, we observe that individuals with severe COVID-19 recognize a smaller number of pressured epitopes. It is important to note that the COVID-19 blood samples were collected in September 2020 when the Wuhan strain was the most prevalent in India. Thus, higher recognition of pressured epitopes by asymptomatic individuals compared to those with severe COVID-19 could possibly imply that the epitopes which initially offered protectivity were under higher immune pressure, resulting in emergence of SARS-CoV-2 variants with mutations in these regions as a means of immune escape.

At the HLA-allele level, we ranked alleles based on the number of SARS-CoV-2 epitopes they bind to, with high affinity, which is used as a predictive measure of CTL response. Based on their occurrence in individuals with various disease states, we assigned a protectivity score to HLA-alleles and epitopes. We shortlisted protective HLA-alleles and epitopes which display a protectivity score larger than a threshold value corresponding to mild and asymptomatic disease states. Recognition of protective epitopes showed strong negative correlation with COVID-19 severity in patients. Among protective HLA-alleles, HLA-B*15:03 which has been shown to be protective against viral infections in previous studies^27^ appeared among the top ranked HLA alleles in our study in terms of CTL response as well as protectivity score. In addition, HLA-B*46:01 did not show high affinity binding with any of the SARS-CoV-2 epitopes in our study, agreeing with previous reports^27^. However, some of the top responding alleles reported in literature were not captured in the top ranked alleles in our study since we considered a large number of indigenous alleles, some of which respond better than the global high frequency alleles documented in literature. We also note that CTL response is a more accurate method of ascertaining HLA allele protectivity than the concept of allele protectivity in clinical samples used in our study since disease outcome and hence protectivity is determined by the HLA genotype as a whole rather than individual HLA alleles. In summary, we notice that the protectivity offered by certain epitopes of SARS-CoV-2 against severe COVID-19 by virtue of an effective CTL response generated in the host, subjects these epitopes to selection pressure.

We acknowledge that many of the results presented here might be influenced by some of the limitations that accompany the approaches undertaken. Some of these include – i) the TAP proteasomal pathway, vesicular trafficking of HLA bound peptides from the ER to the cell surface and absence of TCRs specific to viral epitopes which resemble self-peptides due to thymic negative selection, are further aspects which need to be considered while quantifying CTL responses based on epitope presentation. ii) High affinity binding of certain epitopes could not be validated due to lack of previously available experimental binding affinity studies of indigenous HLA alleles considered in our study iii) COVID-19 is known to present a wide range of immunopathologies in patients^6^ due to which CTL response need not be a major contributing factor in determining disease severity in some cases. Thus, our hypothesis of lower CTL responses in severe disease states might not be applicable to all the COVID-19 patient samples considered in our study. Despite these and the small sample size in each category, a clear trend appears to be there. In the future, our framework allows incorporation of some of these aspects into our analysis once the aforementioned data is made available.

Our current analysis lays emphasis on the role played by host genetic heterogeneity in determining COVID-19 disease outcome and driving evolution of new SARS-CoV-2 variants. The results obtained in our study provide deeper evolutionary insights regarding the prevailing host-pathogen arms race in nature along with potential clinical implications. Identifying pressured epitopes within populations could aid in prediction of possible mutations in future variants of SARS-CoV-2 based on the local selection pressure imposed by highly represented HLA-alleles within the population. Future directions to our study include incorporation of HLA Class-II genotypes and B cell responses in analysing COVID-19 disease susceptibilities at the population level. Large-scale *in silico* analysis of viral and host genomes in recent years has providing a holistic approach towards analysing host-pathogen interactions and ascertaining evolutionary paths adopted by pandemic-or epidemic-causing pathogens.

## Methods

### SARS-CoV-2 protein sequences and HLA Class-I allele data

A total of 11 SARS-CoV-2 variants including 4 Variants of Concern (B.1.1.7 - Alpha, B.1.351 - Beta, P.1 - Gamma, B.1.617.2 - Delta), 4 Variants of Interest (B.1.525 - Eta, B.1.526 - Iota, C.37 - Lambda, B.1.617.1 - Kappa) as listed in the official WHO webpage as of August, 2021 along with 2 other Variants namely, B.1.526.1 and P.2 and the Wuhan strain (A) were considered for our study. Amino acid sequences of 10 proteins, namely - ORF1ab polyprotein, Spike glycoprotein, ORF3a, Envelope protein, Membrane protein, ORF6, ORF7a, ORF8, Nucleocapsid protein and ORF10 were obtained from NCBI Virus^23^ (https://www.ncbi.nlm.nih.gov/labs/virus/vssi/#/).

A total of 1827 HLA Class-I alleles were obtained from 240 ethnic groups with HLA-allele frequency data from the Allele Frequency Net Database^25^ (AFND) (http://www.allelefrequencies.net/). The ethnic groups were shortlisted based on i) documentation of frequencies of all 3 HLA Class I alleles - A, B and C, ii) High resolution HLA allele data with polymorphisms denoted for all the alleles documented.

### Prediction of CTL epitopes

HLA Class I Epitope predictions for the 10 proteins were performed using the NetMHCPan BA 4.1 tool^28^ made available by IEDB (Immune Epitope Database)^24^ (http://tools.iedb.org/mhci/). A comprehensive list of 1827 HLA Class I alleles covered by the 240 ethnic groups were considered for pMHC binding predictions. Length of the predicted epitopes was restricted to 9 amino acids. Predicted 9-mers which bind to a given HLA allele with IC50 < 50 nM were considered as strong binding epitopes. Epitope predictions were performed for all 11 variants of SARS-CoV-2.

### Immune pressure parameters

For the immune pressure plot, 2 parameters were considered for each protein: the protein sequence pairwise distance and the epitope set pairwise distance between each pair of variants. These parameters are defined as follows:

Let V_1_ and V_2_ be the two variants being compared and let P_1_ = [a_1i_]_1≤i≤n_ and P_2_ = [a_2j_]_1≤j≤n_ be the aligned amino acid sequences of a particular protein belonging to the 2 variants with an alignment length n. Let E_1_ = [e_1a_]_1≤a≤p_ and E_2_ = [e_2b_]_1≤b≤q_ be the sets of distinct epitopes of the protein corresponding to the variants V_1_ and V_2_ respectively, where p and q are the respective number of epitopes of the protein.

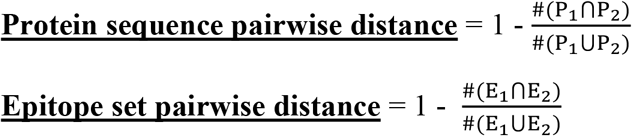

These 2 parameters are calculated for each pair of variants and are represented as points in a 2D scatter plot (Figure 2A, B)

### Shannon Entropy and Epitope Density

For each residue of a protein, 2 further parameters namely, Shannon Entropy and Epitope Density are calculated as follows:

Let P_V_ = [p_i_]_1≤i≤11_ be the set of amino acid sequences for a particular protein across the 11 SARS-CoV-2 variants along with the Wuhan strain considered, where p_i_ = [a_ij_]_1≤j≤n_ for all 1≤ i ≤ 11 is the amino acid sequence of protein p of variant i with an alignment length of n. Let R_j_= [r_ij_]_1≤i≤11_ be the set of unique amino acids across variants for protein p at residue j and 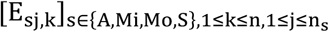 where m_kj_ is the number of variants possessing amino acid a_kj_ at position j.

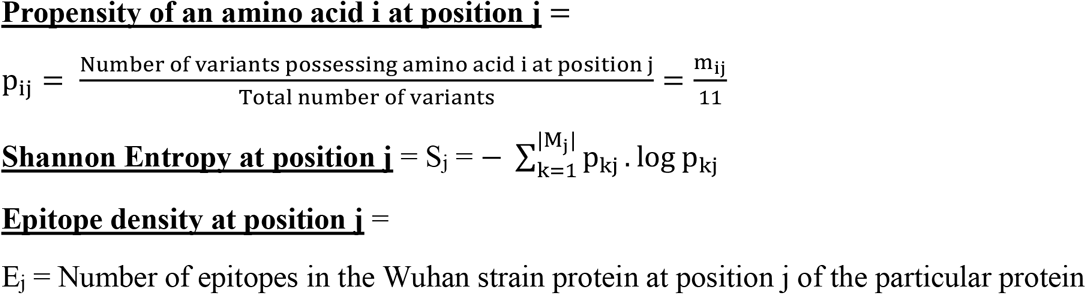

### Support from Literature for the CTL epitopes

Out of 577 experimentally validated epitopes from over 25 publications^29^, 482 high binding epitopes appeared in our study. The total epitope pool of 2022 epitopes in case of the SARS-CoV-2 Wuhan strain could not be covered completely due to binding of some epitopes to indigenous HLA alleles, which have not been validated experimentally in previous studies.

### COVID-19 sample collection

Ethical approval for this study was obtained from the Institutional Ethics Committee at Bangalore Medical College and Research Institute, Bangalore, India (BMCRI/PS/02/2021-21), and IISc (1-26062020), Bangalore, India. Written informed consent was obtained from all study participants before sample collection.

COVID-19 patients were recruited for this study at the hospital upon obtaining informed consent from the participants, during September-December, 2020. The presence of the disease was confirmed by qPCR for viral RNA in nasal and throat swabs of the individuals. Patients who had prior episodes of COVID-19 infection, or had chronic infections such as tuberculosis, HIV or HCV were excluded. A total of 36 Covid-19 patients (median age = 52 + 16, both male and female participants), volunteered their samples for this study. The disease severity classification was performed based on the clinical symptoms presented by the patient at the hospital, which are as follows: Asymptomatic (n=7), Mild (n=9), Moderate (n=10) and Severe (n=10). Whole blood samples were collected in PAXgene Blood RNA tubes and stored at -20°C until further use.

### RNA isolation and HLA Typing

RNA isolation was performed using Paxgene Blood RNA kit (Cat# 762164) following manufacturer protocol. It was then subjected to quantification and quality control, followed by RNA sequencing. 1μg of total RNA from samples with RIN > 5 was used for poly-A mRNA enrichment with NEBNext Poly(A) mRNA Magnetic Isolation Module (NEB #E7490), followed by library preparation with NEBNext Ultra II Directional RNA Library Prep Kit (NEB #E7760) using manufacturer directions. The paired-end RNA sequencing was performed in an Illumina Hiseq 4000 instrument. The library preparation and sequencing was performed by Biokart India Private Limited, Bangalore. The raw sequencing data was subjected to adapter trimming with the tool TrimGalore (version 0.6.6) and Class-I HLAs were predicted from the RNA sequencing reads using the software arcasHLA v0.2.0^30^ with standard settings.

### Epitope and HLA allele protectivity

Protectivity of epitopes and HLA alleles were calculated based on the HLA genotypes corresponding to the clinical samples with varying disease severities.

Let [HG_Aj_]_1≤j≤7_, [HG_Mij_]_1≤j≤9_, [HG_Moj_]_1≤j≤10_ and [HG_Sj_]_1≤j≤10_ be the sets of individuals with asymptomatic, mild, moderate and severe COVID-19 respectively, represented by their HLA-genotypes. Each HLA genotype is comprised of a set of 6 HLA alleles. Since some HLA-alleles are associated with more than one disease severity, the frequency of association of a HLA allele H with a disease severity DS, f_H,DS_ where DS ∈ {A, Mi, Mo, S} is calculated as follows:

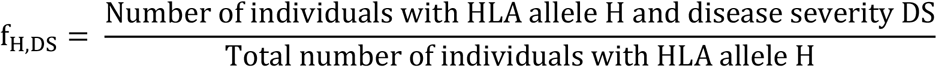

Each HLA genotype is also associated with a corresponding set of distinct epitopes 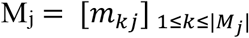 where n is the number of epitopes recognized by the HLA-alleles comprising the genotype and n_s_ is the number of individuals with disease severity s recognizing the epitope. Since epitope sets recognized by individuals with different disease severities overlap in many cases, the frequency of association of an epitope E with a disease severity DS, f_E,DS_ where DS ∈ {A, Mi, Mo, S} is calculated for each epitope as follows:

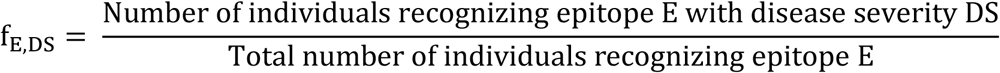

A protectivity value is then assigned for each disease state as follows:

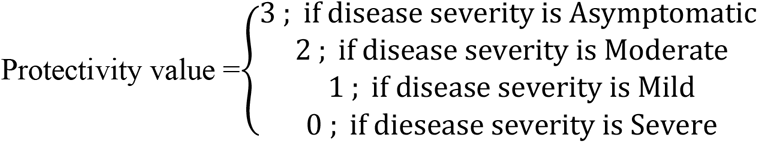

The protectivity score for a HLA-allele H is then calculated as follows:

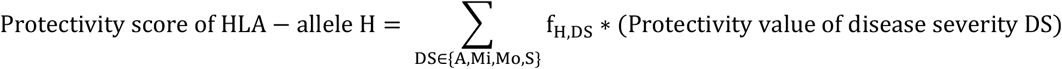

The protectivity score for an epitope E is calculated as follows:

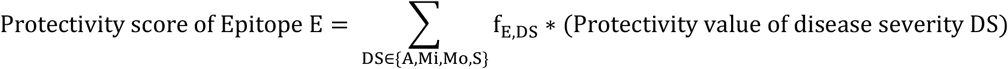

## Data Availability

All data produced in the present study are available upon reasonable request to the authors

https://www.ncbi.nlm.nih.gov/labs/virus/vssi/#/

http://www.allelefrequencies.net/

## Supplementary Material

Supplementary Figures and Tables Supplementary File-1

