## Supplementary Figures and Tables for "Predicted CTL responses from pressured epitopes in SARS-CoV-2 correlate with COVID-19 severity"

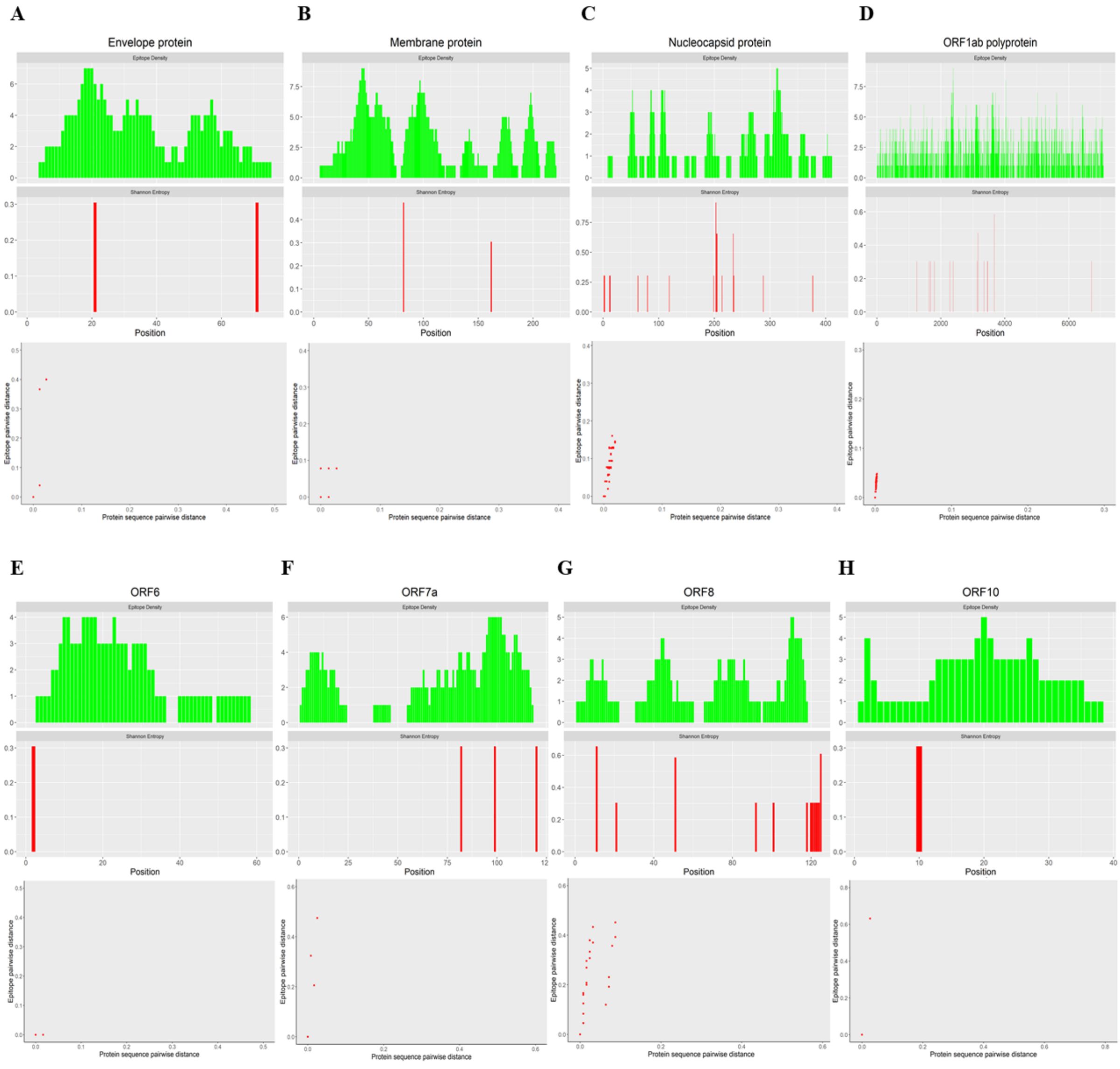


**Supplementary Figure 1:** Analysis of CTL epitopes of (A) Envelope protein, (B) Membrane protein, (C) Nucleocapsid protein, (D) ORF1ab polyprotein, (E) ORF6, (F) ORF7a, (G) ORF8 and (H) ORF10, under immune pressure. Symmetric difference between epitope sets for each pair of variants is plotted against the whole-protein sequence distance (bottom). The points in the scatter plot lie above the diagonal (y=x) for all proteins except Membrane protein and ORF6, indicating higher selection pressure on epitopic regions. Sequence variation in the protein is computed as the Shannon entropy at each residue of the protein (middle - red). Distribution of epitopes within the protein is represented as the epitope density at each residue (middle - green).

| Sl No. | HLA Class-I allele | Number of binding pressured epitopes |
|---|---|---|
| 1 | B*15:62 | 40 |
| 2 | A*02:11 | 35 |
| 3 | B*15:103 | 34 |
| 4 | B*15:132 | 34 |
| 5 | B*15:156 | 34 |
| 6 | B*15:173 | 34 |
| 7 | B*15:03 | 34 |
| 8 | B*15:61 | 33 |
| 9 | A*02:22 | 31 |
| 10 | A*02:104 | 31 |
| 11 | A*02:50 | 31 |
| 12 | A*02:122 | 28 |
| 13 | A*68:23 | 25 |
| 14 | A*02:79 | 24 |
| 15 | A*02:21 | 24 |
| 16 | A*02:61 | 24 |
| 17 | A*02:06 | 24 |
| 18 | A*02:28 | 24 |
| 19 | C*12:19 | 24 |
| 20 | A*02:137 | 24 |
| 21 | A*02:51 | 24 |
| 22 | C*03:40 | 23 |
| 23 | C*03:43 | 23 |
| 24 | A*02:90 | 23 |
| 25 | A*23:04 | 23 |
| 26 | C*03:42 | 23 |
| 27 | A*24:94 | 23 |
| 28 | B*15:25 | 22 |
| 29 | B*35:10 | 22 |
| 30 | B*15:39 | 22 |
| 31 | A*02:76 | 21 |
| 32 | A*02:47 | 21 |
| 33 | A*02:102 | 20 |
| 34 | C*14:02 | 20 |
| 35 | C*14:03 | 20 |
| 36 | C*14:11 | 20 |
| 37 | A*02:115 | 20 |
| 38 | A*02:63 | 20 |
| 39 | C*14:10 | 20 |
| 40 | C*12:20 | 20 |
| 41 | A*02:02 | 20 |
| 42 | A*02:229 | 20 |
| 43 | A*02:186 | 20 |
| 44 | A*02:209 | 20 |
| 45 | B*48:02 | 20 |
| 46 | A*24:23 | 20 |
| 47 | A*24:33 | 20 |
| 48 | A*24:03 | 20 |
| 49 | A*02:131 | 19 |
| 50 | A*68:02 | 19 |
| 51 | A*02:16 | 19 |
| 52 | A*68:44 | 19 |
| 53 | A*68:27 | 19 |
| 54 | B*35:28 | 18 |
| 55 | B*35:41 | 18 |
| 56 | A*02:44 | 18 |
| 57 | C*14:05 | 18 |
| 58 | A*02:158 | 18 |
| 59 | C*03:60 | 17 |
| 60 | A*02:230 | 17 |
| 61 | C*03:02 | 17 |
| 62 | B*15:17 | 17 |
| 63 | A*02:03 | 17 |
| 64 | A*02:264 | 17 |
| 65 | A*24:138 | 17 |
| 66 | C*12:24 | 16 |
| 67 | A*02:48 | 16 |
| 68 | A*02:14 | 16 |
| 69 | B*35:35 | 16 |
| 70 | A*24:10 | 16 |
| 71 | A*24:22 | 16 |
| 72 | A*02:35 | 16 |
| 73 | A*02:167 | 16 |
| 74 | A*02:05 | 16 |
| 75 | A*02:85 | 15 |
| 76 | A*02:177 | 15 |
| 77 | A*02:212 | 15 |
| 78 | A*02:29 | 15 |
| 79 | A*02:96 | 15 |
| 80 | A*02:09 | 15 |
| 81 | A*02:145 | 15 |
| 82 | A*02:01 | 15 |
| 83 | A*02:40 | 15 |
| 84 | B*15:123 | 15 |
| 85 | A*02:121 | 15 |
| 86 | A*02:42 | 15 |
| 87 | A*02:211 | 15 |
| 88 | A*02:157 | 15 |
| 89 | B*35:49 | 15 |
| 90 | A*02:74 | 15 |
| 91 | A*02:118 | 15 |
| 92 | A*02:24 | 15 |
| 93 | A*02:161 | 15 |
| 94 | A*02:221 | 15 |
| 95 | A*02:30 | 15 |
| 96 | A*02:68 | 15 |
| 97 | A*02:93 | 15 |
| 98 | A*02:138 | 15 |
| 99 | A*02:133 | 15 |
| 100 | A*02:16 | 19 |

**Supplementary Table 1**: List of top 100 HLA-alleles exhibiting high affinity binding with pressured CTL epitopes

| Sample No. | HLA genotype | COVID-19 severity |
|---|---|---|
| 1 | A*30:01 \| A*33:03 \| B*15:02 \| B*13:02 \| C*06:02 \| C*08:01 | Severe |
| 2 | A*68:01 \| A*11:01 \| B*40:06 \| B*15:01 \| C*03:03 \| C*14:02 | Severe |
| 3 | A*03:01 \| A*11:01 \| B*15:05 \| B*40:01 \| C*03:03 \| C*03:04 | Mild |
| 4 | A*02:11 \| A*01:01 \| B*35:03 \| B*52:01 \| C*12:02 \| C*12:03 | Mild |
| 5 | A*11:01 \| A*33:03 \| B*52:01 \| B*58:01 \| C*03:02 \| C*12:02 | Mild |
| 6 | A*01:01 \| A*24:02 \| B*07:02 \| B*37:01 \| C*07:02 \| C*06:02 | Mild |
| 7 | A*33:03 \| A*33:03 \| B*13:01 \| B*51:01 \| C*04:03 \| C*14:02 | Mild |
| 8 | A*03:01 \| A*32:01 \| B*35:01 \| B*35:01 \| C*04:01 \| C*04:01 | Mild |
| 9 | A*02:11 \| A*11:01 \| B*40:06 \| B*51:01 \| C*15:02 \| C*07:02 | Mild |
| 10 | A*02:11 \| A*01:01 \| B*51:01 \| B*08:01 \| C*07:02 \| C*14:02 | Mild |
| 11 | A*24:02 \| A*24:02 \| B*44:03 \| B*35:03 \| C*07:06 \| C*12:03 | Moderate |
| 12 | A*11:01 \| A*11:01 \| B*13:01 \| B*52:01 \| C*04:03 \| C*12:02 | Moderate |
| 13 | A*01:01 \| A*01:01 \| B*37:01 \| B*57:01 \| C*06:02 \| C*06:02 | Severe |
| 14 | A*33:03 \| A*26:01 \| B*44:03 \| B*49:01 \| C*07:06 \| C*07:01 | Severe |
| 15 | A*11:01 \| A*24:02 \| B*51:01 \| B*57:01 \| C*07:02 \| C*06:02 | Severe |
| 16 | A*02:11 \| A*02:01 \| B*27:07 \| B*40:06 \| C*15:02 \| C*15:02 | Moderate |
| 17 | A*24:02 \| A*32:01 \| B*15:02 \| B*07:02 \| C*07:02 \| C*08:01 | Moderate |
| 18 | A*24:02 \| A*24:02 \| B*40:06 \| B*52:01 \| C*12:02 \| C*03:04 | Moderate |
| 19 | A*02:11 \| A*02:11 \| B*40:06 \| B*07:06 \| C*15:02 \| C*07:02 | Severe |
| 20 | A*02:11 \| A*01:01 \| B*35:01 \| B*51:01 \| C*16:02 \| C*04:01 | Moderate |
| 21 | A*01:01 \| A*11:01 \| B*49:01 \| B*07:06 \| C*07:01 \| C*07:02 | Severe |
| 22 | A*01:01 \| A*24:02 \| B*52:01 \| B*57:01 \| C*06:02 \| C*12:02 | Moderate |
| 23 | A*30:01 \| A*11:01 \| B*07:02 \| B*13:02 \| C*07:02 \| C*06:02 | Moderate |
| 24 | A*02:11 \| A*02:06 \| B*35:01 \| B*58:01 \| C*03:02 \| C*04:01 | Asymptomatic |
| 25 | A*11:01 \| A*68:01 \| B*40:06 \| B*15:01 \| C*03:03 \| C*14:02 | Asymptomatic |
| 26 | A*01:01 \| A*03:01 \| B*40:06 \| B*50:01 \| C*15:02 \| C*06:02 | Mild |
| 27 | A*33:03 \| A*24:02 \| B*35:01 \| B*58:01 \| C*03:02 \| C*04:01 | Asymptomatic |
| 28 | A*26:01 \| A*24:02 \| B*35:03 \| B*51:01 \| C*04:01 \| C*14:02 | Asymptomatic |
| 29 | A*33:03 \| A*24:02 \| B*35:01 \| B*58:01 \| C*03:02 \| C*04:01 | Asymptomatic |
| 30 | A*03:02 \| A*29:01 \| B*35:03 \| B*07:05 \| C*15:05 \| C*04:01 | Asymptomatic |
| 31 | A*02:11 \| A*01:01 \| B*51:01 \| B*08:01 \| C*07:02 \| C*14:02 | Moderate |
| 32 | A*02:11 \| A*11:01 \| B*40:06 \| B*51:01 \| C*15:02 \| C*07:02 | Moderate |
| 33 | A*33:03 \| A*24:02 \| B*07:02 \| B*07:02 \| C*07:02 \| C*07:02 | Severe |
| 34 | A*01:01 \| A*01:01 \| B*37:01 \| B*57:01 \| C*06:02 \| C*06:02 | Severe |
| 35 | A*24:02 \| A*02:03 \| B*15:02 \| B*38:02 \| C*07:02 \| C*08:01 | Asymptomatic |
| 36 | A*11:01 \| A*24:02 \| B*52:01 \| B*40:01 \| C*12:02 \| C*03:04 | Severe |

**Supplementary Table 2**: List of HLA-genotypes obtained via HLA-typing of blood samples obtained from COVID-19 patients along with the corresponding COVID-19 severity

| Sl No. | Epitope | Protein |
|---|---|---|
| 1 | TANPKTPKY | ORF1ab polyprotein |
| 2 | LGAENSVAY | Spike glycoprotein |
| 3 | FPFTIYSLL | ORF10 |
| 4 | VAVKMFDAY | ORF1ab polyprotein |
| 5 | YINVFAFPF | ORF10 |
| 6 | VFAFPFTIY | ORF10 |
| 7 | STQDLFLPF | Spike glycoprotein |
| 8 | ITVNVLAWL | ORF1ab polyprotein |
| 9 | QWSLFFFLY | ORF1ab polyprotein |
| 10 | VPFWITIAY | ORF1ab polyprotein |
| 11 | SANLAATKM | Spike glycoprotein |
| 12 | QIYKTPPIK | Spike glycoprotein |
| 13 | HTIDGSSGV | ORF3a |
| 14 | TVAYFNMVY | ORF1ab polyprotein |
| 15 | TVNVLAWLY | ORF1ab polyprotein |
| 16 | MYIFFASFY | ORF1ab polyprotein |
| 17 | ASLPFGWLI | ORF3a |
| 18 | YLALYNKYK | ORF1ab polyprotein |
| 19 | GVYYHKNNK | Spike glycoprotein |
| 20 | STVFPPTSF | ORF1ab polyprotein |
| 21 | KATYKPNTW | ORF1ab polyprotein |
| 22 | IQASLPFGW | ORF3a |
| 23 | LPPAYTNSF | Spike glycoprotein |
| 24 | AVKMFDAYV | ORF1ab polyprotein |
| 25 | VMFTPLVPF | ORF1ab polyprotein |

**Supplementary Table 3**: List of pressured-protective CTL epitopes

<table>
<tr><th>HLA Class-I genotype</th></tr>
<tr><td>A*02:03 | A*02:03 | B*07:05 | B*07:05 | C*15:05 | C*15:05</td></tr>
<tr><td>A*02:03 | A*02:03 | B*07:05 | B*38:02 | C*15:05 | C*15:05</td></tr>
<tr><td>A*02:03 | A*02:03 | B*38:02 | B*07:05 | C*15:05 | C*15:05</td></tr>
<tr><td>A*02:03 | A*02:03 | B*38:02 | B*38:02 | C*15:05 | C*15:05</td></tr>
<tr><td>A*02:03 | A*03:02 | B*07:05 | B*07:05 | C*15:05 | C*15:05</td></tr>
<tr><td>A*02:03 | A*03:02 | B*07:05 | B*38:02 | C*15:05 | C*15:05</td></tr>
<tr><td>A*02:03 | A*03:02 | B*38:02 | B*07:05 | C*15:05 | C*15:05</td></tr>
<tr><td>A*02:03 | A*03:02 | B*38:02 | B*38:02 | C*15:05 | C*15:05</td></tr>
<tr><td>A*02:03 | A*29:01 | B*07:05 | B*07:05 | C*15:05 | C*15:05</td></tr>
<tr><td>A*02:03 | A*29:01 | B*07:05 | B*38:02 | C*15:05 | C*15:05</td></tr>
<tr><td>A*02:03 | A*29:01 | B*38:02 | B*07:05 | C*15:05 | C*15:05</td></tr>
<tr><td>A*02:03 | A*29:01 | B*38:02 | B*38:02 | C*15:05 | C*15:05</td></tr>
<tr><td>A*02:03 | A*02:06 | B*07:05 | B*07:05 | C*15:05 | C*15:05</td></tr>
<tr><td>A*02:03 | A*02:06 | B*07:05 | B*38:02 | C*15:05 | C*15:05</td></tr>
<tr><td>A*02:03 | A*02:06 | B*38:02 | B*07:05 | C*15:05 | C*15:05</td></tr>
<tr><td>A*02:03 | A*02:06 | B*38:02 | B*38:02 | C*15:05 | C*15:05</td></tr>
<tr><td>A*03:02 | A*02:03 | B*07:05 | B*07:05 | C*15:05 | C*15:05</td></tr>
<tr><td>A*03:02 | A*02:03 | B*07:05 | B*38:02 | C*15:05 | C*15:05</td></tr>
<tr><td>A*03:02 | A*02:03 | B*38:02 | B*07:05 | C*15:05 | C*15:05</td></tr>
<tr><td>A*03:02 | A*02:03 | B*38:02 | B*38:02 | C*15:05 | C*15:05</td></tr>
<tr><td>A*03:02 | A*03:02 | B*07:05 | B*07:05 | C*15:05 | C*15:05</td></tr>
<tr><td>A*03:02 | A*03:02 | B*07:05 | B*38:02 | C*15:05 | C*15:05</td></tr>
<tr><td>A*03:02 | A*03:02 | B*38:02 | B*07:05 | C*15:05 | C*15:05</td></tr>
<tr><td>A*03:02 | A*03:02 | B*38:02 | B*38:02 | C*15:05 | C*15:05</td></tr>
<tr><td>A*03:02 | A*29:01 | B*07:05 | B*07:05 | C*15:05 | C*15:05</td></tr>
<tr><td>A*03:02 | A*29:01 | B*07:05 | B*38:02 | C*15:05 | C*15:05</td></tr>
<tr><td>A*03:02 | A*29:01 | B*38:02 | B*07:05 | C*15:05 | C*15:05</td></tr>
<tr><td>A*03:02 | A*29:01 | B*38:02 | B*38:02 | C*15:05 | C*15:05</td></tr>
<tr><td>A*03:02 | A*02:06 | B*07:05 | B*07:05 | C*15:05 | C*15:05</td></tr>
<tr><td>A*03:02 | A*02:06 | B*07:05 | B*38:02 | C*15:05 | C*15:05</td></tr>
<tr><td>A*03:02 | A*02:06 | B*38:02 | B*07:05 | C*15:05 | C*15:05</td></tr>
<tr><td>A*03:02 | A*02:06 | B*38:02 | B*38:02 | C*15:05 | C*15:05</td></tr>
<tr><td>A*02:03 | A*02:03 | B*07:05 | B*07:05 | C*15:05 | C*15:05</td></tr>
</table>

<table>
<tr><th>HLA Class-I genotype</th></tr>
<tr><td>A*29:01 | A*02:03 | B*07:05 | B*07:05 | C*15:05 | C*15:05</td></tr>
<tr><td>A*29:01 | A*02:03 | B*07:05 | B*38:02 | C*15:05 | C*15:05</td></tr>
<tr><td>A*29:01 | A*02:03 | B*38:02 | B*07:05 | C*15:05 | C*15:05</td></tr>
<tr><td>A*29:01 | A*02:03 | B*38:02 | B*38:02 | C*15:05 | C*15:05</td></tr>
<tr><td>A*29:01 | A*03:02 | B*07:05 | B*07:05 | C*15:05 | C*15:05</td></tr>
<tr><td>A*29:01 | A*03:02 | B*07:05 | B*38:02 | C*15:05 | C*15:05</td></tr>
<tr><td>A*29:01 | A*03:02 | B*38:02 | B*07:05 | C*15:05 | C*15:05</td></tr>
<tr><td>A*29:01 | A*03:02 | B*38:02 | B*38:02 | C*15:05 | C*15:05</td></tr>
<tr><td>A*29:01 | A*29:01 | B*07:05 | B*07:05 | C*15:05 | C*15:05</td></tr>
<tr><td>A*29:01 | A*29:01 | B*07:05 | B*38:02 | C*15:05 | C*15:05</td></tr>
<tr><td>A*29:01 | A*29:01 | B*38:02 | B*07:05 | C*15:05 | C*15:05</td></tr>
<tr><td>A*29:01 | A*29:01 | B*38:02 | B*38:02 | C*15:05 | C*15:05</td></tr>
<tr><td>A*29:01 | A*02:06 | B*07:05 | B*07:05 | C*15:05 | C*15:05</td></tr>
<tr><td>A*29:01 | A*02:06 | B*07:05 | B*38:02 | C*15:05 | C*15:05</td></tr>
<tr><td>A*29:01 | A*02:06 | B*38:02 | B*07:05 | C*15:05 | C*15:05</td></tr>
<tr><td>A*29:01 | A*02:06 | B*38:02 | B*38:02 | C*15:05 | C*15:05</td></tr>
<tr><td>A*02:06 | A*02:03 | B*07:05 | B*07:05 | C*15:05 | C*15:05</td></tr>
<tr><td>A*02:06 | A*02:03 | B*07:05 | B*38:02 | C*15:05 | C*15:05</td></tr>
<tr><td>A*02:06 | A*02:03 | B*38:02 | B*07:05 | C*15:05 | C*15:05</td></tr>
<tr><td>A*02:06 | A*02:03 | B*38:02 | B*38:02 | C*15:05 | C*15:05</td></tr>
<tr><td>A*02:06 | A*03:02 | B*07:05 | B*07:05 | C*15:05 | C*15:05</td></tr>
<tr><td>A*02:06 | A*03:02 | B*07:05 | B*38:02 | C*15:05 | C*15:05</td></tr>
<tr><td>A*02:06 | A*03:02 | B*38:02 | B*07:05 | C*15:05 | C*15:05</td></tr>
<tr><td>A*02:06 | A*03:02 | B*38:02 | B*38:02 | C*15:05 | C*15:05</td></tr>
<tr><td>A*02:06 | A*29:01 | B*07:05 | B*07:05 | C*15:05 | C*15:05</td></tr>
<tr><td>A*02:06 | A*29:01 | B*07:05 | B*38:02 | C*15:05 | C*15:05</td></tr>
<tr><td>A*02:06 | A*29:01 | B*38:02 | B*07:05 | C*15:05 | C*15:05</td></tr>
<tr><td>A*02:06 | A*29:01 | B*38:02 | B*38:02 | C*15:05 | C*15:05</td></tr>
<tr><td>A*02:06 | A*02:06 | B*07:05 | B*07:05 | C*15:05 | C*15:05</td></tr>
<tr><td>A*02:06 | A*02:06 | B*07:05 | B*38:02 | C*15:05 | C*15:05</td></tr>
<tr><td>A*02:06 | A*02:06 | B*38:02 | B*07:05 | C*15:05 | C*15:05</td></tr>
<tr><td>A*02:06 | A*02:06 | B*38:02 | B*38:02 | C*15:05 | C*15:05</td></tr>
</table>

**Supplementary Table 4**: List of HLA-genotypes offering highest protectivity against severe COVID-19
